# The ImmuneRACE Study: A Prospective Multicohort Study of Immune Response Action to COVID-19 Events with the ImmuneCODE™ Open Access Database

**DOI:** 10.1101/2020.08.17.20175158

**Authors:** Jennifer N. Dines, Thomas J. Manley, Emily Svejnoha, Heidi M. Simmons, Ruth Taniguchi, Mark Klinger, Lance Baldo, Harlan Robins

## Abstract

**Objectives:** The primary aim of this study is to increase our understanding of the adaptive immune response to the SARS-CoV-2 virus by assaying the peripheral immune repertoire for virus-associated T-cell receptors (TCRs). Secondary aims include identification and characterization of SARS-CoV-2–specific B-cell receptors (BCRs) and monoclonal antibodies (mAbs) generated by antibody-producing cells during and after acute infection.

**Trial design:** ImmuneRACE is a prospective, single group, multi-cohort, exploratory study of unselected eligible participants exposed to, infected with, or recovering from SARS-CoV-2.

**Participants:** Approximately 1000 individuals, aged 18 to 89 years and residing in 24 different geographic areas within the United States, will be enrolled, primarily using remote telemedicine technologies. Cohorts will be based on clinical history. Cohort 1 will include participants exposed to SARS-CoV-2 within 2 weeks of study entry. Cohort 2 participants will include those clinically diagnosed or with positive laboratory confirmation of active COVID-19 disease. Cohort 3 will comprise participants previously diagnosed with COVID-19 disease who have been deemed recovered based on two consecutive negative tests, clearance by a healthcare professional, or resolution of symptoms related to COVID-19. All participants must be able to communicate with the investigator and understand and comply with the requirements of the study. Protected populations and those who may not safely participate are not eligible for this study.

**Intervention and comparator:** Blood samples and nasopharyngeal or oropharyngeal swabs will be collected from participants. Nasopharyngeal or oropharyngeal swabs will be collected by inserting a swab into the nose or throat of the participant. Samples will be shipped frozen or transported refrigerated or at room temperature to Adaptive Biotechnologies for processing, including, but not limited to, testing for coronavirus or other respiratory illnesses, DNA extraction, and TCR analysis. The immunoSEQ assay will be conducted using DNA extracted from blood samples. An electronic questionnaire will be administered to collect information pertaining to the participant’s medical history, symptoms, and diagnostic tests performed for COVID-19 disease. Participants will have the option to undergo additional blood draws and questionnaires over a 2-month period. In collaboration with Microsoft, we will use machine learning and artificial intelligence approaches to construct a classifier based on TCR repertoire data designed to accurately distinguish COVID-19–positive cases from unexposed controls (from the ImmuneCODE database of TCR sequences). A rigorous statistical analysis will be performed to validate the classifier.

**Main outcomes:** The main outcomes of this study will include a comparison of disease-specific TCR signatures in patients and controls, identification of the immunodominant antigens that elicit a T-cell response to SARS-CoV-2, risk stratification based on an individual’s immune signature, and determination of immune signatures of patients exposed to SARS-CoV-2 that may allow earlier detection of infection compared to available tests.

**Trial registration:** “ImmuneRACE – Immune Response Action to COVID-19 Events (Protocol ADAP-006, version 1.0; 5/8/2020) is registered with the US National Institutes of Health and can be accessed at ClinicalTrials.gov (NCT04494893).

## I. BACKGROUND

Adaptive Biotechnologies has developed a platform technology for immune profiling called the T-cell receptor βeta-1 (TCRβ1) Assay, also referred to as immunoSEQ^§^ in this protocol. The platform utilizes next-generation sequencing (NGS) to identify rearranged T-cell receptor beta (TCRβ) gene sequences and characterize the abundance of those sequences. This broad platform technology can be used to determine the diversity of the cellular adaptive immune system and track phenotype-associated T-cell clones to query the current state and history of the adaptive immune system.

### Detecting Specific TCRs for Diagnostics^1–3^

Despite the overwhelming number of possible T-cell receptors (TCRs), many TCRs are quite common and repeatedly recur within and between individuals, comprising the public T-cell repertoire^4^. These public TCRs are diagnostically useful because once a TCR has been associated with a particular antigen or disease state, the association is reliable, even when that TCR is seen in a new individual. Using this technique, we have demonstrated that CMV infection can be assayed purely based on the peripheral TCR repertoire^3^.

When specific TCRs associated with particular antigens or disease states are known, scanning the T-cell repertoire for those specific sequences can be clinically useful. The purpose of this research study is to explore the immune system in individuals with coronavirus disease.

## II. PROPOSED RESEARCH

The SARS-CoV-2 virus that causes coronavirus disease (COVID-19) is spreading rapidly throughout the world. Researchers, governments, and biotechnology companies are mobilizing to develop and disseminate diagnostic and therapeutic alternatives to try to curb this global pandemic. The vast majority of these R&D efforts are focused on the RNA of the virus itself, rather than critical information held within the genetics of a patient’s immune response to the virus and the disease patterns that can be inferred through study of the immune response at the population level.

First, we will identify antigens, the parts of the virus that induce a cellular immune response via T cells. We do this by rapidly determining the nearly full set of processed, presented, immunogenic, and immunodominant antigens from a disease. Antigen identification has historically been very challenging for the immunology community, and we are confident that our approach is a significant breakthrough in the field. Next, we use these antigens to identify tens of thousands of T-cell receptors from T cells expanded in response to the virus across the human population. This computational challenge requires the scale of data generation and processing made possible by the Adaptive and Microsoft partnership.

In parallel, we will identify and confirm the TCR signature of the immune response to the SARS-CoV-2 virus. TCR signatures will be identified in DNA extracted from peripheral or whole blood samples from approximately 1000 patients with COVID-19. The immune signature of COVID-19 we expect to identify is a starting point to solve key public health challenges in the present. These samples can either be obtained from newly diagnosed patients or from patients who have already recovered from the virus because the immune response will still be identifiable in the memory compartment.

## III. PURPOSE OF THE STUDY

The primary aim is to improve our understanding of the immune system response in coronavirus diseases by assaying the peripheral immune repertoire of TCRs and secondarily of BCRs, specific to severe acute respiratory syndrome coronavirus 2 (SARS-CoV-2), the virus that causes COVID-19, with the potential to develop better diagnostic tests and treatments.

Adaptive and Microsoft are committed to making TCR repertoire data publicly available to researchers around the world. We hypothesize that by studying the immune response in patients with COVID-19 and making these data public, we can help solve several key challenges plaguing this current pandemic.

### Specific Aims

#### Aim 1. Comparison of disease-specific TCR signatures in patients and controls

We will run immunoSEQ on approximately 1000 patient samples with disease status unblinded. After sequencing these samples, we will quantitatively describe the compartment of the T-cell repertoire specific for the disease of interest. We will then use these data to construct a classifier that accurately distinguishes patients from controls.

#### Aim 2. Identify the immunodominant antigens that elicit a T-cell response to COVID-19

We have developed technology to query the antigen specificity of the T-cell repertoire against hundreds of peptide epitopes in a single blood sample^5^. This technology not only allows us to parse out immunodominant epitopes of a given infection from irrelevant epitopes, but also allows us to determine the TCR sequences of the T cells responding to these immunodominant epitopes. We will use this technology to determine which peptide epitopes derived from the SARS-CoV-2 genome commonly elicit an immunodominant T-cell response in different samples. The TCR sequence data generated from these studies will be used to boost the diagnostic classifier obtained in Aim 1. In addition, the identification of immunodominant epitopes can be disseminated to optimize studies in vaccine design and understanding the relationship between antigen-specific T-cell responses and clinical outcomes in other labs.

#### Aim 3. Risk stratification based on an individual’s immune signature

There is a critical need for a reliable risk stratification test to enable treatment prioritization given the potentially massive number of symptomatic patients. Despite an emerging understanding of the diagnosis of COVID-19, how it spreads, and the death rate, there is currently no available means to predict who needs hospitalization beyond age-associated risk and limited epidemiologically-linked comorbidities. We aim to utilize the immune signature as a way to predict disease severity in individuals by utilizing T-cell sequencing capabilities and machine learning pipelines to identify T cells specific to the virus and assessing the strength of the immune response by the expansion of these T cells in a patient’s blood. By understanding the strength and specificity of a patient’s immune response, which may be temporarily bolstered by recent exposure to other coronaviruses (e.g., common cold) in some people, we may be able to significantly improve the diagnostic and risk stratification processes.

#### Aim 4. Determine whether an immune signature can be detected in individuals exposed to SARS-CoV-2 earlier than currently available tests

Another critical need to contain the spread of SARS-CoV-2 is to determine the false negative rate of the RNA test in asymptomatic people and offer an alternative diagnostic that is more sensitive in this cohort. For example, it is possible that early-stage disease is not picked up by RNA tests because the virus may be isolated to regions such as the lower respiratory cavity that are not assessed by standard testing methods. We aim to determine whether the immune response can be used to detect the virus from a simple blood test, thereby providing a more sensitive test in asymptomatic people, even if the virus itself is not directly detectable in the upper respiratory region.

#### Secondary Aim

##### Aim 5. Explore whether additional research assays could potential identify and/or confirm antigenic binding

As a secondary aim, Adaptive will perform exploratory research with additional sequencing-based research assays to profile the adaptive immune system, including, but not limited to, TCR pairSEQ^**^^6^ and B-cell receptor (BCR) pairSEQ^††^. These assays use a combinatorial method for pairing TCR alpha and beta chain sequences and BCR heavy and light chain sequences. The output is a large set of full-length, paired BCR or TCR sequences, which allows reconstruction of a functional antibody or TCR. We regularly utilize the results of our pairSEQ assays to identify and/or confirm antigenic binding. For the case of BCR pairSEQ, the resulting antibody sequences could potentially have therapeutic value for imparting passive immunity.

## IV. STUDY POPULATION

### Participants

Approximately 1000 individuals, between the ages of 18 and 89, who reside within the United States will be prospectively ascertained. Blood samples and nose or throat swabs will be collected at affiliated sites or with mobile phlebotomy. These samples will be shipped frozen or transported refrigerated or at room temperature to Adaptive Biotechnologies for processing, including, but not limited to, DNA extraction and analysis. Minors, pregnant women, incarcerated individuals, mentally-disabled persons, and wards of the state will be excluded to prevent any risk to vulnerable populations. The selection of participants will be equitable per 45 CFR 111(a). The inclusion and exclusion criteria for the three cohorts are included below.

### Cohort 1. Exposed to coronavirus disease

#### Inclusion criteria

Participants must satisfy the following criteria to be enrolled in the study:

i. Individuals exposed to someone with a confirmed diagnosis of coronavirus disease within 2 weeks of enrollment (or at the discretion of the investigator)
ii. Male and female participants of any race and ethnicity between 18 to 89 years of age (inclusive) at the time of enrolling in the study
iii. Must be able to communicate with the investigator and understand and comply with the requirements of the study

#### Exclusion Criteria

The presence of any of the following will exclude a participant from enrollment:

i. Individuals who have not been exposed to a person with a confirmed diagnosis of coronavirus disease within 2 weeks of enrollment (or at the discretion of the investigator)
ii. Protected populations, including minors, pregnant women, incarcerated individuals, mentally-disabled persons, and wards of the state
iii. Any significant condition, laboratory abnormality, or psychiatric illness that would prevent the participant from safely participating in the study
iv. Donated more than 500 cc or 1 pint of blood in the past 60 days prior to the blood draw (at the discretion of the investigator)

### Cohort 2. Active coronavirus disease

#### Inclusion criteria

Participants must satisfy the following criteria to be enrolled in the study:

i. Individuals with a diagnosis of coronavirus disease, either by:

a. Clinical diagnosis made by a medical professional, or
b. Positive laboratory test, including but not limited to naso- or oropharyngeal swab (or at the discretion of the investigator)
ii. Male and female participants of any race and ethnicity between 18 and 89 years of age (inclusive) at the time of enrolling in the study
iii. Must be able to communicate with the investigator and understand and comply with the requirements of the study

#### Exclusion Criteria

The presence of any of the following will exclude a participant from enrollment:

i. Individuals without a diagnosis of coronavirus disease
ii. Protected populations, including minors, pregnant women, incarcerated individuals, mentally-disabled persons, and wards of the state
iii. Any significant condition, laboratory abnormality, or psychiatric illness that would prevent the participant from safely participating in the study
iv. Donated more than 500 cc or 1 pint of blood in the past 60 days prior to the blood draw (at the discretion of the investigator)

### Cohort 3. Recovered from coronavirus disease

#### Inclusion criteria

Participants must satisfy the following criteria to be enrolled in the study:

i. Individuals previously diagnosed with coronavirus disease and cleared from active infection based on:

a. Testing negative on two consecutive naso- or oropharyngeal swab tests following initial diagnosis, or
b. Clearance by a healthcare professional or public health authority, or
c. Resolution of symptoms related to COVID-19 (or at the discretion of the investigator)
ii. Male and female participants of any race and ethnicity between 18 and 89 years of age (inclusive) at the time of enrolling in the study
iii. Must be able to communicate with the investigator and understand and comply with the requirements of the study

## III. Exclusion Criteria

The presence of any of the following will exclude a participant from enrollment:

i. Individuals without a previous diagnosis of coronavirus disease at the discretion of the investigator
ii. Protected populations, including minors, pregnant women, incarcerated individuals, mentally-disabled persons, and wards of the state
iii. Any significant condition, laboratory abnormality, or psychiatric illness that would prevent the participant from safely participating in the study
iv. Donated more than 500 cc or 1 pint of blood in the past 60 days prior to the blood draw (at the discretion of the investigator)

## V. METHODS AND PROCEDURES

### Sample and Data Collection Plan

#### Blood Draws and Sample Processing

Standard blood draws will be performed via routine venipuncture during either routine clinical care of the participants or scheduled remote phlebotomy appointments for the research study. Whole blood will be collected in volumes of approximately 10–60 mL (∼1–6 × 10 mL tubes). Within 72 hours of being drawn, all blood samples will be shipped frozen (at −20 to −80 °C) or transported refrigerated (at 2 to 8°C) to Adaptive Biotechnologies for further testing. Blood will undergo DNA extraction, and the immunoSEQ Assay will be performed. ImmunoSEQ results will quantitatively describe the compartment of the T-cell repertoire specific for the disease of interest compared to controls. Additional research assays may be applied.

Depending on the needs of the study, Adaptive will coordinate phlebotomist blood draws that will be transported the same day at room temperature for processing. These samples must be processed and cryopreserved within 6 hours of the sample blood draw. Both PBMCs and serum will be processed and frozen. Samples prioritized for these procedures will include, but are not limited to, those from individuals with a confirmed diagnosis of coronavirus disease and reporting symptoms for at least two weeks prior to blood draw.

#### Nose or Throat Swabs

Nasopharyngeal or oropharyngeal swabs will be collected by inserting a swab into the nose or throat of the participant. Collected samples will be shipped frozen at −80 °C to Adaptive within 1 month of collection or stored at 4 °C and transported to Adaptive within 72 hours. All samples will be stored at −80°C until further testing is performed. Testing may include, but is not limited to, clinically available tests for coronavirus disease or other respiratory illnesses.

#### Electronic Questionnaire

An electronic questionnaire will be administered in this study to collect information pertaining to the participant’s medical history, symptoms, and diagnostic tests performed for coronavirus disease. This information will be used in the analysis to better understand and interpret the test results. Responses will be stored and labeled with an alphanumeric code. All information will be de-identified in accordance with HIPAA standards.

As part of this study, medical records, with the participant’s consent, may be accessed to confirm medical information pertaining the participant’s medical history, symptoms, and diagnostic tests performed for coronavirus disease.

#### Optional Participation

Participants will have the option to undergo up to four additional blood draws and four additional questionnaires about the participant’s symptoms and medical information relating to coronavirus disease over a 2-month time period. Each additional blood draw will be approximately 10–60 mL (1–6 × 10-mL tube(s)). Optional blood draws are at the discretion of the investigator and dependent on the needs of the study.

Additional tests to diagnose or support the diagnosis may be run on samples from participants enrolled in all cohorts. This possibility is explicitly stated in the consenting information sheet.

### Shipping

Batches of samples can be transported to Adaptive by courier. All sample shipments will follow regulations for UN 3373 Biological Substance, Category B. Shipment notification and the electronic manifest should be provided before sample receipt at Adaptive.

Contact: Emily Svejnoha +1 (206) 693-2032 Address:

ATTN: BSM Adaptive Biotechnologies 1551 Eastlake Ave E, Suite 200 Seattle, WA 98102

### Metadata Summary

For each sample the following metadata will be included, but is not limited to:

i. Basic demographics (age at collection, gender, ethnicity/race)
ii. Symptoms associated with coronavirus disease and at time of blood draw
iii. Required hospitalizations relating to coronavirus disease
iv. Date and city/state of diagnosis of coronavirus disease
v. Test results for coronavirus disease
vi. Other comorbid conditions that may impact results and interpretation (for example, asthma, chronic lung disease, autoimmune conditions)
vii. Medications that may impact results and interpretation

### Data Analysis

Data analysis will be performed by Adaptive Biotechnologies scientists using customized molecular assays and software designed to capture detailed information about the immune repertoire. In collaboration with Microsoft, machine learning and artificial intelligence (AI) will be used to construct a classifier that accurately distinguishes cases from controls based on TCR repertoire analysis. A rigorous statistical analysis will be performed.

For the secondary aim, TCR pairSEQ and/or BCR pairSEQ assay analysis will be performed. These analyses are based on a combinatorial method for pairing TCR alpha and beta chain sequences and BCR heavy and light chain sequences, allowing reconstruction of a functional antibody or TCR.

This study is considered minimal risk and does not require additional data monitoring measures.

### Immunodominant peptide analysis

Cryopreserved PBMC will be used for the assay, which will link TCR sequences to T-cell specificity of peptides derived from the SARS-CoV-2 genome as described by Klinger et al.^5^. Briefly, memory T cells from SARS-CoV-2–exposed individuals will be incubated with epitopes derived from the entire genome of SARS-CoV-2. SARS-CoV-2–specific memory T cells will be isolated and sequenced. The TCR sequences of the memory response will be used to broaden the diagnostic signal obtained by repertoire analysis alone. In addition, SARS-CoV-2 antigenic epitopes that are found to be immunogenic in participants who have cleared the infection will be identified and shared with the community to facilitate further studies targeting these epitopes.

### Data Storage and Confidentiality

DNA extracted from the samples will be stored at −20 to −80 °C until the sample is exhausted. Samples and associated clinical information will be de-identified prior to being transferred or upon arrival to Adaptive Biotechnologies. The samples will be stored and tracked by research technicians using a laboratory information management system (LIMS). The samples may be used for research assays in areas beyond the disease of interest at the conclusion of this study.

To the extent that results of this research study are presented at meetings or in publications, the identities of participants will not be included in the information that is disclosed. In addition, records related to this study may be reviewed during a regulatory inspection.

## VI. RISK/BENEFIT ASSESSMENT

Risk Category: Minimal

### Potential Risk - Physical

Physical risks associated with drawing blood include: soreness at the site of puncture, bruising, and in the rare case, infection of the blood draw site, fainting, nerve, or tendon damage.

Physical risks associated with a nose or throat swab include: a very small risk of bleeding when swabbing the inside of the nose or throat (mucosal membranes).

### Mitigating strategies to minimize risk

Standard procedures will be used to minimize discomfort and the chance of infection resulting from venipuncture, nasal, or throat swabs.

### Potential Risk – Loss of Confidentiality

There may be a risk of loss of confidentiality of the protected health information of the participants.

### Mitigating strategies to minimize risk

All data and blood samples from participants will be de-identified at Adaptive in accordance with the HIPAA Safe Harbor Method. In addition, staff sign confidentiality agreements as conditions of their employment and are trained annually on the HIPAA Privacy and Security Rule. Delegated research staff will be able to view information about the participant, the code given their samples, and the resulting data derived from the analysis. Therefore, appropriate data security measures will be in place to prevent unauthorized access to individually identifiable data.

### Benefits

No direct benefit exists for study participants.

Participants are made aware that allowing the study to use their blood samples and analyze their DNA may help develop better lab tests and lead to a better scientific understanding of coronavirus disease.

In addition, de-identified data will be shared with the global research community with the potential of developing better tests and treatment for coronavirus disease.

The participants are informed that they will not receive individual results from this study. Moreover, we will not provide the results to their doctors or put them in their medical records.

### Alternatives to Participation

Options include not participating or participating in other research studies.

## VII. PARTICIPANT IDENTIFICATION, RECRUITMENT AND CONSENT

### Method of Participant Identification and Recruitment

Prospective participants will be identified and recruited by, but not limited to, clinicians and other health care personnel at participating sites, online and printed marketing material, and radio and television advertisements. The identification and recruitment of participants will protect privacy and be free of undue influence. All material will be submitted to the IRB for approval.

### Process of Consent

Eligible individuals will be given an information sheet and will electronically consent to participate in the study. We are requesting a waiver of signed consent given:

- The research involves no more than minimal risk
- The waiver of informed consent will not adversely affect the rights and welfare of the participant
- It is not practicable to conduct the research without the waiver or alteration
- Whenever appropriate, participants will be provided with additional pertinent information after their participation

Participants who wish to participate in this study will demonstrate consent by clicking “ACCEPT” to an information sheet that describes the study. They will not be able to participate if they do not click “ACCEPT”. Additional information about the study will be available to participants as FAQ. Please reference the FAQ and eConsent information sheet for further details. There will be no elements of coercion or undue influence. Consent will be obtained via an eConsent process and/or through a research coordinator or other appropriate study staff when applicable. Documentation of consent and contact information for those who wish to be contacted in future studies will be obtained and stored on a secure, password-protected server. If eConsent is not available or handwritten signatures are required, a paper consent will be used; the participant will demonstrate consent by signing at the bottom of the information consent sheet that describes the study. For certain instances in which a wet ink signature is required, a HIPAA form to be shared with the provider network will be issued for authorization of release of medical records.

### Costs to the Participant

Adaptive Biotechnologies, the Sponsor, or study partners (Microsoft, LabCorp/Covance, and Illumina), will pay for procedures associated with the study and necessary follow-up. Participants will not incur any costs nor will insurance be billed for research procedures in this study.

### Payment for Participation

Participants will be given a $50 gift card for their participation in the study. Participants must complete the blood draw, nasal or oral swab, and study questionnaire prior to receiving the gift card.

Participants will be provided an additional $50 gift card after completing each additional blood draw and questionnaire about their symptoms and medical information relating to COVID-19.

## VIII. SPONSOR

Adaptive, or study partners (Microsoft, LabCorp/Covance and Illumina), will sponsor the entirety of this study and all related costs, including resource needs to curate metadata.

## Data Availability

Data and updates related to this clinical trial protocol will be curated on ClinicalTrials.gov where the trial has been registered as NCT04494893. Through a collaboration with Microsoft, TCR sequence data resulting from the primary aims of this study will be made publicly available through the open access database ImmuneCODE at www.immunecode.com.

## Declarations

### Ethics approval

The ImmuneRACE Study has been approved by the Western Institutional Review Board (reference number 1-1281891-1) on March 17th, 2020.

### Authors’ contributions

All authors contributed to the drafting and editing of this manuscript. All authors approved the final manuscript. JND contributed to protocol development, study design, study medical lead and manuscript review; TJM contributed to development and review of manuscript content; ES contributed to drafting study materials including protocol and ICF; HMS contributed to manuscript preparation; RT contributed to protocol edits; MK contributed to MIRA panel conception, design and execution (excluding actual lab work) plus analyses; LB contributed to protocol development, study execution and development and review of manuscript content; HR contributed to development and review of manuscript content.

## Acknowledgements

We thank Mark Ellis and Melanie Styers for providing editorial support.

## Footnotes

§ For Research Use Only. Not for use in diagnostic procedures

** For Research Use Only. Not for use in diagnostic procedures

†† For Research Use Only. Not for use in diagnostic procedures

